# Individual brain regulation as learned via neurofeedback is related to affective changes in adolescents with autism spectrum disorder

**DOI:** 10.1101/2022.05.24.22275449

**Authors:** Manfred Klöbl, Karin Prillinger, Robert Diehm, Kamer Doganay, Rupert Lanzenberger, Luise Poustka, Paul Plener, Lilian Konicar

**Author notes:** Correspondence to: Lilian Konicar, Department of Child and Adolescent Psychiatry, Medical University of Vienna.

## Abstract

**Background:** Even though emotions often play a role in neurofeedback (NF) regulation strategies, investigations of the relationship between the induced neuronal changes and improvements in affective domains are scare in electroencephalography-based studies. We thus extend the findings of the first study on slow cortical potential (SCP) NF in autism spectrum disorder (ASD) by relating affective changes to whole-brain activity at rest and during regulation.

**Methods:** Forty-one male adolescents with ASD were scanned at rest using functional magnetic resonance imaging before and after half of them underwent NF training and half received treatment as usual. Furthermore, parents reported on affective characteristics at both times. The NF group had to alternatingly produce negative and positive SCP during training and was additionally scanned while applying their developed regulation strategies.

**Results:** We did not find significant treatment group-by-time interactions in affective or resting-state measures. However, we found increases of resting activity in the anterior cingulate cortex and right inferior temporal gyrus as well as improvements in affective characteristics over both groups. Activation corresponding to SCP differentiation in these regions correlated with the affective improvements. A further correlation was found for Rolandic operculum activation corresponding to positive SCP shifts. There were no significant correlations with the respective achieved SCP regulation during NF training.

**Conclusion:** SCP NF in ASD did not lead to superior improvements in neuronal or affective functioning compared to treatment as usual. However, the affective changes might be related to the individual strategies and their corresponding activation patterns as indicated by significant correlations on the whole-brain level but not the achieved SCP regulation.

**Trial registration:** This clinical trial was registered at drks.de (DRKS00012339) on 20^th^ April, 2017.

## Background

Autism spectrum disorder (ASD) constitutes a pervasive neurodevelopmental disorder which has its onset during childhood and comprises deficits in communication and interaction abilities as well as atypical, repetitive behavior (1). A variety of therapeutic approaches exists, each focusing on different symptoms (2, 3).

A particular family of treatment approaches based on a system- and network-level understanding of ASD comprises the application of non-invasive neuromodulation techniques (4). Especially transcranial direct current (5-7) and magnetic stimulation (8-10) have been explored in the past. In contrast, neurofeedback (NF) is a neuromodulatory intervention without external stimulation.

When undergoing NF, patients train to gain volitional control over certain characteristics of their brain activity, aiming to improve aberrant patterns specific to their condition. While reviews on the application of electroencephalography-based (EEG) NF for the treatment of attention-deficit / hyperactivity disorder (ADHD) have yielded mixed results (11, 12), there is a particular recommendation for slow cortical potentials (SCPs) NF (13).

SCP NF has been successfully applied for the treatment of ADHD in children (14-16), adolescents (17) and adults (18). These very slow EEG fluctuations (typically below 1 Hz) are related to the excitability threshold of the upper cortical layers, where SCP negativity corresponds to increased and SCP positivity to decreased cortical excitability (19). SCPs further constitute suspected contributors to the blood-oxygenation-level-dependent (BOLD) signal measured in functional magnetic resonance imaging (fMRI) (20, 21), which is now also explored as application modality for NF in ASD (22, 23). However, the utility of NF for the treatment of ASD in general has also been questioned (24).

This criticism is based on the high comorbidity of ASD with ADHD with estimates ranging from 37 to 85% (25) rendering it difficult to assign symptom improvements to either of the disorders in isolation. However, previous research using EEG NF in ADHD and ASD often focused on attention deficits (11, 26) while studies on the emotional and empathic components are largely missing. The recently conducted first study on SCP NF in ASD reported positive effects on ASD-specific symptomatology (27) but also showed the complex influence of the ADHD-related problems regarding attention, hyperactivity and impulsivity (28). Overall, transdiagnostic similarities between ASD and ADHD encompass deficits in emotion regulation, emotion recognition, attention, cognitive flexibility, inhibition, reward processing, working memory, organization and planning (29, 30).

Waddington, Hartman (31) identified four classes of emotion recognition impairments in children and adolescents with and without diagnoses of ASD and/or ADHD. Subsequent polygenic risk score analyses revealed that higher scores for ASD were surprisingly related to faster visual emotion recognition (32). Furthermore, a class defined by impulsive and imprecise visual emotion recognition was associated with lower genetic risk of ASD. On the contrary, slower emotion recognition was reported in adults with ASD or ADHD, related to mind wandering in ADHD but slower processing in ASD (33). Another transdiagnostic analysis in preschool children revealed an increased probability for moderate emotion regulation and executive functioning deficits given an ASD or ADHD diagnosis, but for high emotion regulation deficits in ADHD and high executive functioning deficit in ASD (34).

Regarding empathic capabilities, individuals with ASD showed deficits in cognitive empathy but not affective empathy (35, 36). Reports of empathic abilities may further differ between objective and subjective assessments (37). Interpretation of these results in synopsis is undoubtedly complex, which is indicative of the fact that emotion regulation research in ASD is still in its infancy (38). However, the importance and efficacy of therapeutic approaches incorporating emotion regulation training for ASD are already evident (39).

On a neuronal level, resting-state (RS) functional connectivity was long believed to be characteristically altered in ASD, presenting with local over- and long-range under-connectivity but this view has been challenged by more recent studies (see Mohammad-Rezazadeh, Frohlich (40) for a critical review). Moreover, it was shown that even the same regionally specific measures of functional connectivity were not reproducible across research sites (41). However, the average absolute global connectivity, which can be interpreted as a measure of the linear activation dependency across the brain, was found to be lower in children and adolescents with ASD compared to typically developing controls (42). This might indicate global, rather than local alterations.

Besides functional connectivity, differences between individuals with and without ASD were found in measures of brain activity at rest. A meta-analysis combining results from studies using regional homogeneity (ReHo; a measure sensitive to brain activity and local connectivity), amplitude of low-frequency fluctuations (ALFF; activity within a certain frequency band) and cerebral blood flow reported robust over-activation in language-related and motor areas as well as under-activation in the default mode network (43). Moreover, a large-scale, multi-center study reported two clusters in the brain showing abnormalities in RS connectivity and activity across several measures (44): The first cluster reaches from the left posterior insula to the operculum with decreases in voxel-matched homotopic connectivity (i.e., symmetric connectivity), ReHo and degree centrality (a measure of global connectedness). The second cluster is located at the right dorsal superior frontal cortex and shows increases in fractional ALFF (fALFF; an amplitude-normalized version of ALFF), ReHo and degree centrality.

Even though SCPs are a common NF target, only few studies investigated the neural processes accompanying SCP self-regulation using fMRI. Hinterberger, Veit (45) showed that during transfer runs (training runs without feedback for decoupling regulation success from the feedback procedure) SCP negativity is related to wide-spread activation in fMRI. In turn, SCP positivity was associated with widespread deactivation. It was further shown in patients with epilepsy that deactivation during transfer runs only occurred for individuals who successfully learned to self-induce SCP positivity (46).

In order to gain further knowledge on emotion regulation in ASD, we investigated potential treatment-induced changes in several affective characteristics on a subjective level via parental questionnaires. As neuronal outcome measures at rest, we employed three models previously related to alterations in brain connectivity and activity of individuals with ASD (42-44). First, the percent amplitude of fluctuation (PerAF) model was used for assessing resting brain activity (47) constituting a less artifact-prone and easier to interpret derivative of (f)ALFF (43, 44). Second, average brain-wide connectivity was assessed via global functional connectivity (GFC; (48)), a continuous and thus more sensitive alternative to degree centrality (44). Third, ReHo (49, 50) was assessed as measure of local activity and connectivity (43, 44), placing itself conceptually between PerAF and GFC. Finally, based on suggested links between SCPs and BOLD signal (21, 45), we conducted a brain regulation task in the MRI scanner, in which the subjects had to apply the regulation strategies learned, and investigated potential relationships between whole-brain activation corresponding to SCP regulation and affective changes.

## Methods

### Experimental Design

Participants were randomly allocated to either 24 sessions of SCP NF (n_1_ = 21) or treatment as usual (TAU; n_2_ = 20). SCP feedback was calculated from the fronto-central electrode located according to the extended 10-20 EEG system and presented on a screen as graphical object of the participants’ choice. The participants’ goal was to gain volitional control over their brain activity, moving the object up or down via changes in SCP positivity or negativity (see Konicar, Radev (27) for details on the NF training protocol, EEG artifact correction, feedback presentation, etc.). In order to assess neuronal and subjective changes, fMRI and psychometric data were acquired before the first and after the last treatment session in both groups.

### Participants

Potential male adolescent (12-17 years) participants with a diagnosis of ASD (according to the German version of the “Autism Diagnostic Interview – Revised” (ADI-R) (51) and the “Autism Diagnostic Observation Schedule” (ADOS-2) (52) were recruited and invited to a pre-screening. The further inclusion criteria were: right-handedness and an IQ above 70 (53, 54). The recruitment was restricted to male adolescents due to the prevalence of ASD in this population. Participants were excluded in case of relevant psychiatric, neurological or internal conditions (head injuries, major axis I diagnosis of psychosis, obsessive-compulsive disorder, severe motor or vocal tics, Tourette syndrome, severe depression with suicidality) or MRI contraindications. Previous NF experience and current participation in pharmacological studies were not allowed. Concomitant psychosocial and pharmacological treatments were permitted if kept constant throughout the study participation.

### Psychometric assessment

We used the Emotion Regulation Checklist (ERC (55)) and the Griffith Empathy Measure (GEM (56)) to gather parent reports of the participants’ development regarding their affective and empathic abilities. The emotion regulation (ER) and lability / negativity (LN) subscales of the ERC were analyzed separately, with the former quantifying expression and self-awareness of emotions as well as empathy, and the latter mood lability and anger dysregulation. For the GEM, we concentrated on the cognitive empathy (CE) subscale, since this ability was shown to be diminished in adolescents with ASD in contrast to affective empathy (35).

Since the frequent co-occurrence of ADHD constitutes the major point of discussion to the application of NF to patients with ASD, the “Diagnostic System for Psychiatric Disease in Children and Adolescent, parent-rated version 2” for ADHD (DISYPS-II (57)) was used to quantify the respective characteristics (hereafter referred to as “ADHD score”). The ADHD score was calculated, age-corrected and transformed to the “standard nine” (stanine) score.

### FMRI acquisitions

We positioned the subjects in the MRI scanner and fixated their heads using foam cushions. RS data was acquired as the first functional scan to avoid potential task-related carryover effects. The subjects were instructed to lie with their eyes open, look at a crosshair, let their mind wander and not to think of anything in particular. The recording took 8 minutes.

In the SCP neurofeedback group only, after completing the training, a short brain regulation task was recorded in 2 min 42 s. This was done in order to investigate activation corresponding to the application of the SCP regulation strategies on a whole-brain level. The visual cues for up- and down-regulation were the same as for the SCP transfer run: A triangle pointing upwards (“Up” condition) indicated application of the strategy developed to induce negative SCP shifts and a triangle pointing downwards (“Down” condition) indicated application of the strategy developed to produce positive SCP shifts. The conditions were presented five times each for 8 s in a pseudo-randomized order interleaved with baselines of the same duration. Since the baseline in the SCP transfer run was only 2 s long and had no visual indicator, we added a crosshair of the same size and color scheme as the triangles for the fMRI run. There was no indication of the currently achieved regulation and, contrary to the SCP transfer run, no reward given after regulation trials.

Measurements were performed on a Siemens Magnetom Prisma 3 T machine (Siemens, Erlangen, Germany) with the same sequence as in Moessnang, Schäfer (58) due to the previous successful application in a comparable population: echo / repetition time = 30 / 2000 ms, 3 mm isotropic resolution (+25% gap), 33 slices with 64 × 64 voxels (field of view = 192 × 192 × 123 mm), bandwidth = 2365 Hz/Px. Prospective acquisition correction (PACE) was used for online motion correction.

### FMRI preprocessing

Unless mentioned otherwise, preprocessing was conducted using Statistical Parametric Mapping, version 12 (SPM12). In a combined first step, physiological artifacts were reduced using PESTICA (59) and slice-wise motion correction was performed with SLOMOCO (60). This advanced motion correction approach was taken since adolescents show markedly more in-scanner motion than adults, for which most procedures are optimized. Slice-timing was corrected to the temporally middle slice. All acquisitions per paradigm were realigned together for each participant. A population-specific normalization template was created using the CerebroMatic toolbox (61) with spatially adaptive non-local means and hidden Markov random field filtering for increased homogeneity over the whole age range. Affine regularization was performed to the standard ICBM template for European brains with parameters downscaled by a factor 10 for better local fitting. This was done since some brains showed unreasonable inflation without regularization. Thus, tissue distributions are age-adjusted but the localization of the regions approximately in standard space. The original voxel size was used for reslicing (62). The BrainWavelet toolbox (63) was employed for non-linear artifact correction providing additional mitigation of motion and other types of artifacts. The “threshold” parameter was set to “15” due to the application to unsmoothed data and “chsearch” to “harsh” to be more sensitive towards slow artifacts. The data was finally smoothed with a Gaussian kernel with full width at half maximum (FWHM) of 3 times the voxel size.

### FMRI modelling

An adapted CompCor approach (64, 65) and the Fristion-24 model (66) were utilized for reduction of any residual physiological or movement-related artifacts in the brain regulation task and RS data. The latter was further band-limited to 0.01-0.10 Hz using frequency regressors (67). Based on the filtered RS time series, three voxelwise models were set up: PerAF to quantify brain activity, GFC for brain-wide connectivity and ReHo for local activity and connectivity. Since the calculation of ReHo leads to spatial smoothing, this model was applied to the unsmoothed and filtered data. Afterwards, the ReHo maps were smoothed with a FWHM of 2 times the voxel size achieving smoothness similar to PerAF and GFC. Finally, ReHo and GFC were Fisher z-transformed before group analysis.

The brain regulation task was modeled using the 1st-level module in SPM12. The conditions (“Up” / “Down”) were used as regressors and the CompCor and Friston-24 time series as nuisance signals. In addition, the equivalent of SCP differentiation between changes in negativity and positivity was calculated as the difference between the “Up” and “Down” conditions (henceforth “fMRI differentiation”). The autocorrelation model was set to “FAST” (68).

### Statistical inference

Questionnaire data was analyzed using linear mixed effects models (LMEs). Interactions between “treatment group”, “time” (factors) and “ADHD baseline score” (covariate) were analyzed and dropped if non-significant. The LMEs also included participants as random intercepts.

Whole-brain inference was conducted using the 2nd-level module in SPM12. “Treatment group” by “time” and “time” effects were analyzed in one model per RS measure. Contrasts for within-group effects were further estimated in case of non-significant interactions. Correlations of regulation direction-specific activation and fMRI differentiation with the ERC and GEM score changes were of interest for the brain regulation task. Family-wise error-corrected (FWE) results are reported at the cluster- (primary threshold p ≤ 0.001) or peak-level. Influences of ADHD were controlled for with the pre-training ADHD scores as covariate.

Associations between the RS findings and questionnaire score changes over time were investigated on an exploratory basis using partial correlation (Pearson or Spearman, depending on a visual check of the distributions, corrected for treatment group; median cluster values were extracted using the MarsBaR toolbox 0.44).

In case of significant correlations between regulation activation in the brain regulation task and questionnaire score changes, the latter were subsequently correlated with the average amount of SCP regulation corresponding to the activation contrast achieved during the third and last quarter (six days each) of NF training. These periods showed the strongest regulation or were closest to the second MRI session (27). This way, we checked whether NF training success was related to the score changes (c.f., Heinrich, Gevensleben (69)).

All tests (including neuroimaging models (70)) were two-sided and multiplicity-corrected to p ≤ 0.05. To correct for the number of questionnaire scales / RS models / brain regulation task correlations, we employed an in-house developed algorithm based on the dependency-adjusted D/AP approach (71) (see supplement for implementation).

## Results

Complete RS data was available from 36 and brain regulation task data from 20 participants (Table 1; three subjects did not participate in the second session for personal reasons, two had to be excluded due to compromised data quality and one due to missing compliance).

**Table 1:** Overview of the psychometric and behavioral data.

| Score | Neurofeedback<br>pre | Neurofeedback<br>post | Treatment as<br>usual pre | Treatment as<br>usual post | p-value |
| --- | --- | --- | --- | --- | --- |
| N | 21 | scores: 21<br>resting-state: 19<br>brain regulation<br>task: 20 | 20 | scores: 20<br>resting-state:<br>17 |  |
| Age [years] (at first<br>magnetic resonance<br>imaging session) | 14.44 ± 1.90 |  | 15.01 ± 1.59 |  | 0.2779 <sup>1</sup> |
| Emotion Regulation<br>Checklist: emotion<br>regulation subscale | 21.19 ± 3.98 | 23.05 ± 3.23 | 20.45 ± 5.00 | 21.15 ± 3.59 | time: 0.0419 <sup>2</sup> |
| Emotion Regulation<br>Checklist: lability /<br>negativity subscale | 33.71 ± 5.83 | 30.76 ± 5.34 | 32.00 ± 7.06 | 31.65 ± 6.43 | time: 0.0506 <sup>2</sup><br>ADHD score:<br>0.0014 <sup>2</sup> |
| Griffith Empathy<br>Measure: Cognitive<br>empathy subscale | -1.00 ± 9.85 | 5.33 ± 9.80 | 0.60 ± 9.20 | 1.20 ± 8.89 | time: 0.0601 <sup>2</sup> |
| mean total<br>displacement | 2.49 ± 0.88 | 2.12 ± 0.46 | 2.01 ± 0.41 | 2.16 ± 0.42 | interaction:<br>0.0024 <sup>2</sup><br>group:<br>0.0145 <sup>2</sup> |

Pre- and post-treatment scores are given as mean ± standard deviation. ADHD: attention-deficit / hyperactivity disorder. <sup>1</sup> two-sample t-test, <sup>2</sup> linear mixed effects model.

The results of the SCP regulation training and ASD core symptoms longitudinally quantified via the Social Responsiveness Scale (72) are described in Konicar, Radev (27). Further information on the exact training protocol can also be found in Konicar, Radev (27). A detailed analysis of the comorbid ADHD symptoms and their influence on attention and expectancy measures via the contingent negative variation (CNV) in the EEG signal is presented in Prillinger, Radev (28).

### Psychometric and behavioral analysis

No significant interaction effects were found for any psychometric scale. The ER subscale (higher is better) of the ERC showed a significant symptom improvement over both groups, whereas the LN (lower is better) and GEM CE (higher is better) subscales barely fell short of statistical significance. The LN subscale positively correlated with the pre-training ADHD score.

Since the PerAF results indicated a potential relationship with in-scanner movement, the mean of the total displacement output of the SLOMOCO step of RS preprocessing was additionally analyzed via LME to uncover potential further relationships with psychopathology. There was a significant time by group interaction where the SCP NF group showed significantly higher pre-training movement. Total displacement did not correlate with the ADHD score over both scans (Spearman ρ = −0.05, p = 0.6790). No multiplicity correction was applied in the total displacement analysis.

### Resting-state models

Over both groups, an increase in RS activity was found in the ventral anterior cingulate cortex (ACC). Within the TAU group, a further increase was detected in the right precentral gyrus. Another cluster of increased resting activity over both groups reaching into the medial and inferior temporal gyrus (ITG) did not survive correction for the number of models. Since, upon visual inspection (see Figure 1), especially the result in the precentral gyrus might be biased by baseline differences, LMEs of the median values extracted from the clusters were run (treatment group, time and their interaction as factors, the ADHD score as covariate, random intercept per participant). These confirmed the time effect for the ACC (p = 0.0006) and temporal gyrus (p = 5.4E-6) and indicated a baseline difference (p = 0.0326), time (p = 3.5E-5) and interaction effects (p = 0.0020; all uncorrected) for the precentral gyrus. The ADHD score had no significant influence in either model. Details are presented in the upper section of Table 2 and left column of Figure 1.

**Table 2:**
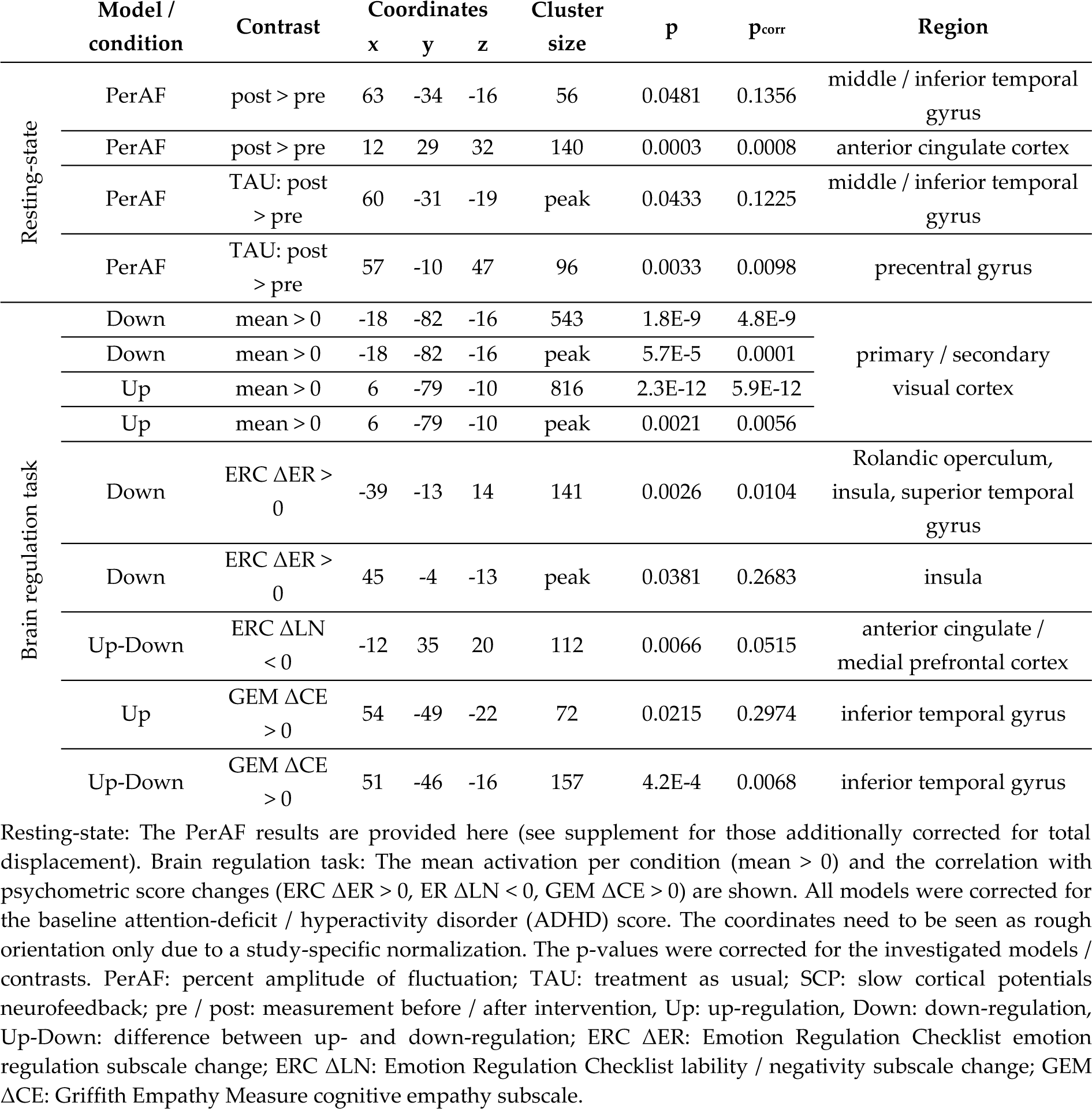
Results of the whole-brain resting-state analysis and brain regulation task.

|  | Model / condition | Contrast | Coordinates |  |  | Cluster size | p | p<sub>corr</sub> | Region |
| --- | --- | --- | --- | --- | --- | --- | --- | --- | --- |
|  |  |  | x | y | z |  |  |  |  |
| Resting-state | PerAF | post > pre | 63 | -34 | -16 | 56 | 0.0481 | 0.1356 | middle / inferior temporal gyrus |
|  | PerAF | post > pre | 12 | 29 | 32 | 140 | 0.0003 | 0.0008 | anterior cingulate cortex |
|  | PerAF | TAU: post > pre | 60 | -31 | -19 | peak | 0.0433 | 0.1225 | middle / inferior temporal gyrus |
|  | PerAF | TAU: post > pre | 57 | -10 | 47 | 96 | 0.0033 | 0.0098 | precentral gyrus |
| Brain regulation task | Down | mean > 0 | -18 | -82 | -16 | 543 | 1.8E-9 | 4.8E-9 | primary / secondary visual cortex |
|  | Down | mean > 0 | -18 | -82 | -16 | peak | 5.7E-5 | 0.0001 |  |
|  | Up | mean > 0 | 6 | -79 | -10 | 816 | 2.3E-12 | 5.9E-12 |  |
|  | Up | mean > 0 | 6 | -79 | -10 | peak | 0.0021 | 0.0056 |  |
| | Down | ERC ΔER > 0 | -39 | -13 | 14 | 141 | 0.0026 | 0.0104 | Rolandic operculum, insula, superior temporal gyrus |
| | Down | ERC ΔER > 0 | 45 | -4 | -13 | peak | 0.0381 | 0.2683 | insula |
| | Up-Down | ERC ΔLN < 0 | -12 | 35 | 20 | 112 | 0.0066 | 0.0515 | anterior cingulate / medial prefrontal cortex |
| | Up | GEM ΔCE > 0 | 54 | -49 | -22 | 72 | 0.0215 | 0.2974 | inferior temporal gyrus |
| | Up-Down | GEM ΔCE > 0 | 51 | -46 | -16 | 157 | 4.2E-4 | 0.0068 | inferior temporal gyrus |

Resting-state: The PerAF results are provided here (see supplement for those additionally corrected for total displacement). Brain regulation task: The mean activation per condition (mean > 0) and the correlation with psychometric score changes (ERC ΔER > 0, ER ΔLN < 0, GEM ΔCE > 0) are shown. All models were corrected for the baseline attention-deficit / hyperactivity disorder (ADHD) score. The coordinates need to be seen as rough orientation only due to a study-specific normalization. The $p$ -values were corrected for the investigated models / contrasts. PerAF: percent amplitude of fluctuation; TAU: treatment as usual; SCP: slow cortical potentials neurofeedback; pre / post: measurement before / after intervention, Up: up-regulation, Down: down-regulation, Up-Down: difference between up- and down-regulation; ERC ΔER: Emotion Regulation Checklist emotion regulation subscale change; ERC ΔLN: Emotion Regulation Checklist lability / negativity subscale change; GEM ΔCE: Griffith Empathy Measure cognitive empathy subscale.

**Figure 1:**
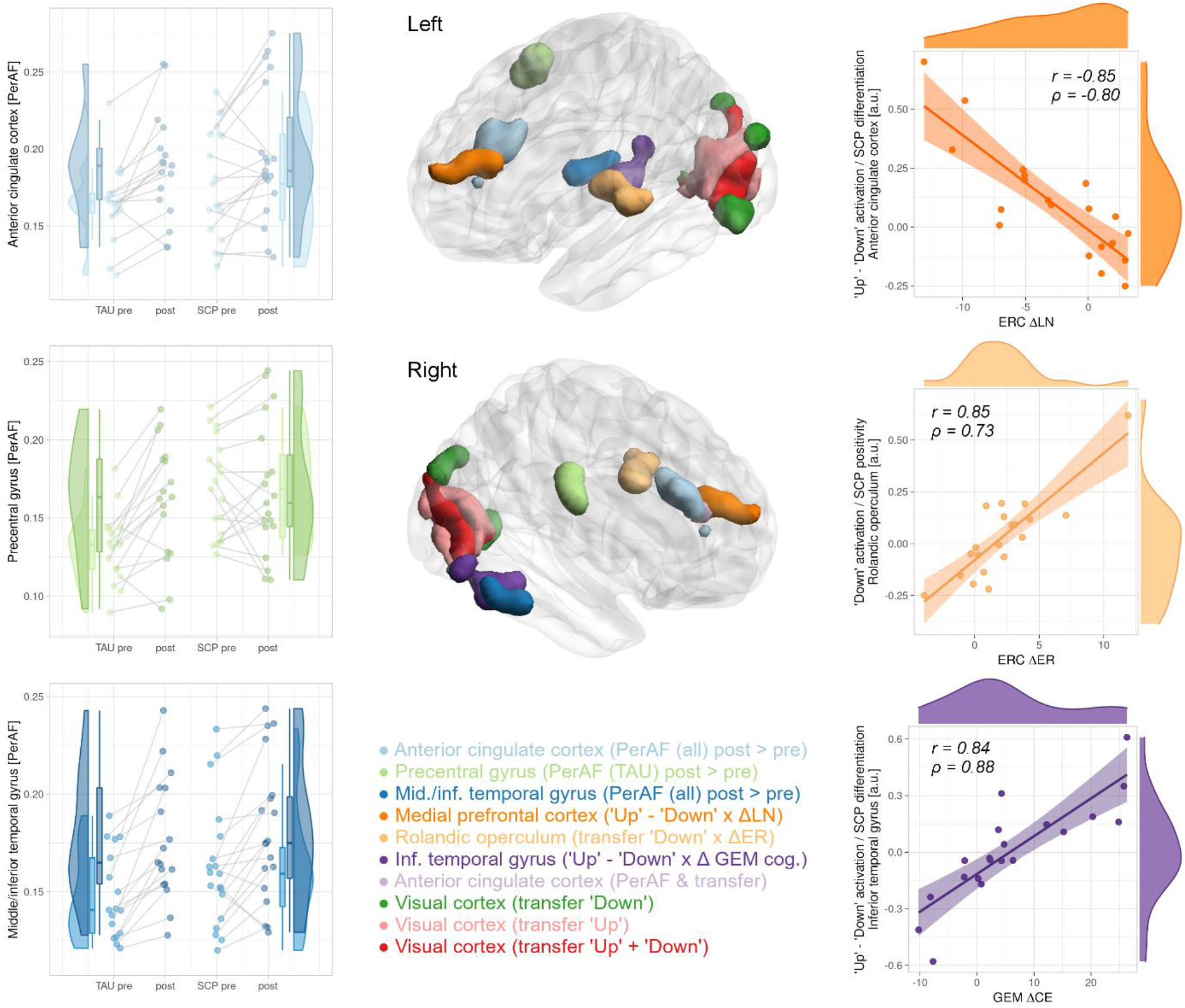
Results of the resting brain activity and brain regulation task analyses. The increase in the percent amplitude of fluctuations (PerAF) of the anterior cingulate cortex over both, the slow cortical potentials (SCP) neurofeedback and treatment as usual (TAU) group, is visualized in the top-left diagram. The middle-left diagram shows a PerAF increase in the precentral gyrus of the TAU and generally higher values in the SCP group. The bottom-left plot shows the PerAF changes over both groups in the temporal gyrus with stronger increases in the TAU group. The right scatter plots show the correlation of the brain regulation task (Pearson’s r corresponding to the parametric analysis as implemented in Statistical Parametric Mapping and Spearman’s ρ due to potential outliers): The difference between the “Up” and “Down” conditions (i.e., fMRI differentiation) was anticorrelated with changes on the lability / negativity (ERC ΔLN) subscale of the Emotion Regulation Checklist. The SCP positivity condition correlated with changes on the emotion regulation (ERC ΔER) subscale. Changes on the Griffith Empathy Measure cognitive empathy (GEM ΔCE) subscale again significantly correlated with fMRI differentiation. The whole-brain 3D models were created using BrainNet Viewer 1.7 (73).

Since precentral gyrus activation might be indicative of movement, the PerAF values of each region were Pearson partially correlated (corrected for measurement, treatment group and their interaction) with the mean total displacement (thresholded at 1.5 times the interquartile range due to potential outliers) and the pre- and post-training ADHD scores on an exploratory basis. Indeed, significant relationships with total displacement were found for all results (r = [0.24, 0.49], p = [0.0479, 1.6E-5]; all p-values for the ADHD score > 0.45). The thresholded mean total displacement values were then used as additional covariate in a repeated analysis. All results survived this control for in-scanner movement with one further cluster for the TAU group in the triangular gyrus (not significant after multiplicity adjustment; see **Fehler! Verweisquelle konnte nicht gefunden werden**., upper section). Given the homogeneous effect on all investigated regions, the influence of total displacement was also investigated on a whole-brain level, resulting in weak but widespread patterns of correlations and anti-correlations (**Fehler! Verweisquelle konnte nicht gefunden werden**.).

Exploratory analyses revealed correlations between changes in the LN subscale and the middle / ITG cluster from both groups (Spearman ρ = 0.41, p = 0.0133). The GFC and ReHo models yielded no significant effects.

### Brain regulation task

During the “Up” and “Down” conditions (corresponding to the induction of negative and positive SCP shifts, respectively), primary and secondary visual areas were activated. No significant difference was found between the two conditions (i.e., no significant fMRI differentiation). Rolandic operculum activation (stretching into the anterior insula and the Heschl gyrus) during the “Down” condition significantly correlated with increases in ER after training. FMRI differentiation in the ITG showed a significant correlation with changes in CE. A further cluster barely missing corrected significance was detected for the correlation of higher fMRI differentiation in the ventral ACC and improvements in LN. A last finding in the right insula did not survive correction for the number of correlations. The baseline ADHD score had no significant influence on any model. For details, see the lower section of Table 2 and right column of Figure 1.

The only notable correlation was found for the change in CE and the average differentiation achieved during the last quarter of the NF training (Spearman: ρ = 0.35, p = 0.1184; all other |ρ| < 0.15, p > 0.53) but was not significant.

## Discussion

In this work, we used fMRI to assess the resting brain activity of adolescents with ASD and collected parental reports of their affective functioning before and after receiving SCP NF or TAU. In addition, the SCP NF group performed a brain regulation task in the scanner after undergoing NF training, where the subjects had to apply their regulation strategies. Using the whole-brain regulation data, we found relationships between the activation during regulation and improvements in different affective domains.

### Affective symptom improvements and the potential influence of regulation strategies

Changes in resting activity and affective improvements over both groups without any interaction effects indicate unspecific positive effects of SCP NF and TAU. Furthermore, while localized correlations between brain activation during regulation and improvements on all investigated affective scales point towards an influence of NF, these improvements did not significantly correlate with SCP regulation (i.e., the SCP shifts produced in the same direction for which the correlations with fMRI activation were detected). Heinrich, Gevensleben (69) likewise concluded universal improvements of emotional and behavioral self-regulation after successfully applying SCP NF in ADHD without finding correlations of training outcomes and symptom improvements. A possible interpretation of these findings is to ascribe the affective improvements not to the achieved SCP regulation but the application of the regulation strategies. The correlation between fMRI differentiation and activation during the “Down” condition with affective improvements despite missing significant effects outside of the visual cortex (or in general for fMRI differentiation) could indeed point towards highly individual activation patterns and strategies. In line with this speculation, Hasslinger, D’Agostini Souto (74) identified emotional strategies as a common regulation approach in SCP NF when treating ADHD. Using the same classifications, strategies from the emotional domain were also frequently reported in this study (see supplement of (27)). Furthermore, ER is known to often be a key factor in the development of successful regulation strategies (75, 76). It, however, should be noted that any comparison to NF training outcomes strongly depends on the definition of learning and regulation success.

Regarding the origin of the correlation between fMRI differentiation in the right ITG and improvements in CE the support of the experimenters needs consideration. Empathic comments after negative performance feedback were shown to decreased negative feelings (77). The participants might have unintentionally related verbal positive reinforcement to their regulation strategies ultimately resulting in CE improvements. In a comparable scenario, the experimenters’ empathy was concluded to be a potential driving factor of subjective improvements in the sham group of a NF study in primary insomnia (78). While the weaker correlation of the “Up” condition alone appear to support this hypothesis, the stronger effect for the fMRI differentiation is probably the best argument against such an additional unintentional effect.

Also the TAU group might have improved due to the supportive clinical counseling received, leading to no significant therapeutic advantage of SCP NF. The positive correlation between the LN subscale and the ADHD baseline score corroborates the well-documented emotion dysregulation in ADHD (79-81). The stronger reduction in head movement (total displacement) in the SCP group, constitutes a potential unspecific training effect. The requirement to sit still for a longer time over repeated NF sessions could have led to the reduced in-scanner movement in the SCP group only.

### The anterior cingulate cortex and emotional negativity

Similar to improvements on the psychometric level, we found an increase in ACC resting activity quantified as PerAF over both groups. Early and replicated neuroimaging evidence for structural and functional alterations in the ACC of patients with ASD was found in smaller gray matter volume and decreased metabolic rate (82, 83). This structural anomaly might appear as lower resting activity following volumetric normalization. A negative correlation between the ADOS and ACC fALFF was found to be a distinctive feature of children diagnosed with “pervasive developmental disorder – not otherwise specified” (84). One may assume that symptomatically stronger subgroups lack the correlation between ACC resting activity and ASD symptoms due to a ceiling effect. A lack of activation in the ACC was also found for a stroop task in ADHD and related to the symptoms of inattention and impulsivity (85), which are likely shared between ADHD and ASD (86).

Beyond changes in PerAF, the ventral part of the ACC also showed a negative correlation between fMRI differentiation in the brain regulation task and the change in the LN subscale of the ERC extending into the medial prefrontal cortex. The amount of SCP differentiation was used as a primary parameter for the analysis of the NF training success as it indicates the participants’ ability to differentiate between the regulation directions (16, 27). The PerAF model, being a measure of amplitude fluctuation and revealing partly overlapping changes in the ACC, might also be interpreted as a measure of fMRI “differentiation” at rest. However, the different sampling time in SCP and fMRI investigations as well as the BOLD signal containing influences of neural activity of all frequency bands might be the reasons for the missing correlation between the PerAF ACC cluster and the LN subscale changes.

The ACC is known to be related to repetitive behavior in ASD (87) but also to cognitive inflexibility in depression (88, 89). The latter is potentially also represented in the LN subscale (90), providing further evidence that behavioral flexibility might be reflected in neuronal flexibility of the ACC.

### The inferior temporal gyrus and cognitive empathy

General as well as regionally specific alterations in temporal lobe structures related to ASD are well-known (91, 92). In a classification tree approach using white and gray matter volumes of various temporal regions, smaller gray matter volume of the right ITG was related to a higher probability of an ASD diagnosis (93). Reduced gray matter volume of the left ITG was also detected in children with low-functioning ASD (94) and related to communication skills (95). As for the ACC, smaller gray matter volume in the ITG might be related to less activity after structural normalization, relating our finding of an increase after NF training to compensatory processes. In ASD, reduced resting activity estimated via ReHo was also recently reported in the left middle temporal gyrus (96). While no correlation with symptom severity was found by the authors, we found a positive relationship of PerAF changes in the affected cluster with changes in emotional negativity (LN subscale). Since this would imply a worsening in emotional negativity accompanying an increase in ITG activity, this correlation is in contradiction with our other findings.

Besides the temporal lobe results found at rest, which did not survive correction for the number of models, we also identified a positive correlation between fMRI differentiation in the right ITG and improvements in CE. Support for laterality of this correlation as well as specificity for cognitive compared to affective empathy is gained from findings in unilateral mesial temporal lobe epilepsy (97). Furthermore, the right ITG was related to personal as well as impersonal emotional imagery whereas it was only impersonal for the left ITG (98). This could imply that the right ITG is involved when one tries to understand another person’s emotions, i.e., displays CE. Preston, Bechara (98) further related reduced neuronal recruitment in a region between the ITG and fusiform gyrus to a lack of CE.

### The Rolandic operculum and emotion regulation

The correlation between the “Down” condition (SCP positivity) and the increase in the ER score mostly covers the posterior Rolandic operculum reaching into the insula and superior temporal gyrus. The Rolandic operculum is involved in processing of visceral signals (99), language encoding (100) and emotion processing (101). Further, reduced activation was found in the left Rolandic Operculum of adults with ASD compared to neurotypical individuals regarding speech (102). However, the Rolandic Operculum showed increased activation during emotion induction with happy compared to sad music (103). The same results were previously obtained for children with low-functioning autism and age-matched controls using speech recordings of their parents and each child’s favorite song containing vocals (104).

These findings suggest two possible conclusions related to NF: First, our reported activation related to improvements on the ER subscale was evoked by self-induced positive mood possibly in combination with sound imagination. Second, less severe language deficits facilitated the development of strategies involving inner speech, which had a positive impact on ER. On the role of SCP positivity in particular, we can only make an indirect assumption: Better SCP differentiation was previously associated with less relaxation when trying to produce negative SCP shifts (105), suggesting that the potentially induced positive mood is a side effect of more relaxation when trying to produce positive shifts. This could again be seen as support for the idea that the individual regulation strategies are responsible for the correlations between brain activation and affective improvements. In accordance with this idea, the self-reported frequency of cognitive reappraisal strategies for ER correlated with the within-run training progress in upregulating the left ventrolateral prefrontal cortex in a previous fMRI NF study (106).

### The precentral gyrus at rest

Lastly, we found an increase in resting activity in the right motor cortex of the TAU group, which was, however, biased by a baseline difference. The unspecific correlations to the amount of in-scanner (head) motion, the fact that motion decreased in the SCP NF but PerAF increases in the TAU group and the effect surviving a correction for motion on group-level (in addition to subject-level) speak against movement-related motor activity as sole cause. Altered connectivity of motor regions (107, 108) as well as motor impairments (109) are known in ASD. Higher visuomotor impairment was associated with increased ALFF in the precentral cortex, among other regions (110). Another study reported increases in gray matter volume in the pre- and postcentral gyri of children with ASD only when no ADHD comorbidity was present (111). In a large sample of children and young adults with ADHD, a reduction in gray matter volume was found in the precentral gyrus and several other regions (112). Assuming that the baseline difference in precentral gyrus PerAF is reflecting more deficits / higher symptom severity in pre-treatment measures in the SCP NF group (i.e., increased movement, higher SRS scores), the observed increase in the TAU group cannot be seen as a positive therapeutic effect. This interpretation would also be in line with the previous report of increased ALFF (110) and our decision to correct for baseline ADHD symptom severity.

### Limitations

Some results barely missed statistical significance after multiplicity adjustment but were deemed relevant in relation to others and thus further discussed. Despite correcting for in-scanner head movement far beyond the standard procedure in multiple steps, residual artifacts are likely present in the data, as can be concluded from the correlation with the RS models (see **Fehler! Verweisquelle konnte nicht gefunden werden**.). However, since the PerAF findings survived an additional control for the quantified motion on group-level, in-scanner movements unlikely are the cause of our findings. A potential approach to further reduce head motion at rest beyond fixation and post-hoc motion correction might be the low-demand video “Inscapes” (113). The brain regulation data could only be reasonably acquired in the NF group and after the training, so all conclusions drawn from the data are necessarily purely correlational. We also kept the acquisition as short as possible expecting increasing movement and decreasing attention throughout each session.

During the brain regulation task, it was not possible to check whether participants really apply the strategies learned. Concurrent EEG recordings were not possible but would have been of little use in this case since, on average, our subjects did not gain control over the SCP signal in the absence of feedback (27).

### Conclusion

Increases in resting activity in regions known to be affected in ASD as well as improvements in several affective domains over both treatment groups indicate unspecific positive effects. In addition, the affective improvements correlated with the activation corresponding to SCP regulation during a brain regulation task in comparable regions. These were however largely unrelated to the achieved degree of SCP regulation during NF training. Besides corroborating the role of regional alterations and affective functioning in ASD, our findings might suggest that what actually leads to affective symptom improvements is the application of distinct NF regulation strategies, rather than SCP NF itself. Future research is needed to clarify the distinct influences of SCP positivity and negativity as well as the role of individual regulation strategies.

## Data Availability

All data produced in the present study are available upon reasonable request to the authors.

## List of abbreviation

(f)ALFF: (fractional) amplitude of low-frequency fluctuations
ACC: anterior cingulate4 cortex
ADHD: attention-deficit / hyperactivity disorder
ADI-R: Autism Diagnostic Interview – Revised
ADOS-2: Autism Diagnostic Observation Schedule, version 2
ASD: autism spectrum disorder
BOLD: blood-oxygenation-level-dependent
CE: cognitive empathy subscale of the GEM
CNV: contingent negative variation
DISYPS-II: Diagnostic System for Psychiatric Disease in Children and Adolescent, parent-rated version 2
EEG: electroencephalography
ER: emotion regulation subscale of the ERC
ERC: Emotion Regulation Checklist
FEW: family-wise error
fMRI: functional magnetic resonance imaging
FWHM: full with at half maximum
GEM: Griffith Empathy Measure
GFC: global functional connectivity
ITG: inferior temporal gyrus
LME: linear mixed effect
LN: lability / negativity subscale of the ERC
NF: neurofeedback
PACE: prospective acquisition correction
PerAF: percent amplitude of fluctuations
ReHo: regional homogeneity
RS: resting-state
SCP: slow cortical potential
SPM12: Statistical Parametric Mapping, version 12
TAU: treatment as usual

## Declarations

### Ethics approval and consent to participate

The overall clinical trial and all parts presented here were conducted in accordance with the Declaration of Helsinki and the good scientific practice guidelines of the Medical University of Vienna. The project was approved by the institutional review board (EK-Nr.: 1850/2015). Informed assent was given by the participants and informed consent was provided by the parents / legal guardian.

### Consent for publication

Not applicable.

### Availability of data and materials

Due to reasons of data protection, the preprocessed data is available only upon reasonable request to the corresponding author.

### Authors’ contributions

LK, LP and RL conceptualized the study. LK acquired the funding. LK, RL and MK planned the MRI measurements. MK conducted the MRI measurements with support from LK, KP, RD and KD. KP, RD, KD and LK performed the NF training and monitored the collection of the questionnaire data. MK analyzed the data presented in this manuscript and compiled the original draft. KP, LK and RL preformed the administrative tasks for this study. PP and RL provided the resources for this study. PP, LP and RL supervised the study. All authors read, critically revised and approved the final manuscript.

## Acknowledgements

We are grateful towards our participants and their parents and the students of the ABC BRAIN LAB, who supported us during this study. Furthermore, we would like to thank the psychologists, psychiatrists and therapists, social workers and hospital staff of the Department of Child and Adolescent psychiatry of the Medical University of Vienna.

