## supplement for "Individual brain regulation as learned via neurofeedback is related to affective changes in adolescents with autism spectrum disorder"

**Implementation of the multiplicity correction algorithm**

A custom multiplicity correction approach was developed due to the need for an ad-hoc procedure that takes dependencies between hypotheses into account when random permutation testing is not feasible (e.g., in case of large amounts of data or heterogeneous hypotheses). The idea is based on the Dubey / Armitage & Parmar method (D/AP) discussed in Sankoh, Huque (1), which estimates the dependencies between multiple endpoints via correlations. There, for the p-value $p_k$ corresponding to the $k$-th out of $K$ endpoints, the adjusted p-value $p_{ka}$ is calculated as

$$p_{ka} = 1 - (1 - p_k)^{m_k}$$

with

$$m_k = K^{1-r_{.k}}$$

and

$$r_{.k} = \frac{1}{K-1}\sum_{j \neq k}^{K} r_{jk}$$

$m_k$ can be seen as estimate of the effective number of independent hypotheses from the perspective of the $k$-th endpoint. $r_{.k}$ is the average correlation of this endpoint with all others. The related $R^2$-adjustment (RSA) uses the $R_k^2$ value conditional on all other endpoints instead of the average correlation, which constitutes the amount of shared variance.

The newly implemented algorithm is based on the following deliberations:

- The false-positive rate (FPR) of the D/AP method increases with increasing correlations between the endpoints but decreases again for very high ones.
- RSA displays a similar but less steep profile. For two endpoints, the explained variance $R^2$ can be estimated as squared correlation.
- Pearson correlations can be expressed as the cosine of the angle between the standardized vectors spanned by the endpoint data.
- The cosines of small angles are more similar than those of larger angles. This behavior is inverse to the initial FPR escalation of the D/AP method.
- Considering all endpoints as tested simultaneously, would make a common estimate of the effective number of independent endpoints more reasonable than separate ones.
- If endpoints also show negative correlations, this leads to an underestimation of the dependencies.

Thus:

- Using a similarity measure that is more sensitive to variations at small dependencies might compensate for the escalation of the FPR. Using $1 - \sin(\alpha)$ instead of $\cos(\alpha)$ fulfills this requirement.
- $\sin(\alpha)$ can easily be calculated from $\cos(\alpha)$ via the trigonometric Pythagorean identity: $1 = \sin^2(\alpha) + \cos^2(\alpha)$.
- Hence, the new sine similarity (SiSi) measure, can be derived from correlation as follows: If $r = \cos(\alpha)$, then $r_{SiSi} = 1 - \sin(\alpha) = 1 - \sqrt{1 - \cos^2(\alpha)} = 1 - \sqrt{1 - r^2}$

- The average of the absolute correlations between all simultaneously tested endpoints will be used instead of separate ones.

The formulae above are therefore adapted as follows:

$$p_{ka} = 1 - (1 - p_k)^m$$

with

$$m = K^{1-r_{SiSi}} = K^{1-(1-\sqrt{1-r^2})} = K^{\sqrt{1-r^2}}$$

and

$$r = \frac{2}{K(K-1)} \sum_{i \neq j}^{K} |r_{ij}|$$

This preserves a geometric interpretation of the similarity metric and also offers a statistical one: If $r^2$ is an estimate of the average shared variance between the endpoints, $\sqrt{1-r^2}$ is an estimate of the correlation between endpoints explaining the missing variance.

A comparison of the FPRs between three approaches to multiplicity correction is given in Table S1: Synthetic multivariate correlated normal distributions were created for Monte Carlo simulations of 10000 datasets, comprising 100 points each. Two-sample t-tests of 50 vs. 50 data points were run in order to estimate the FPR for each setting. Sidak implicitly uses the same formulae as DA/P but assumes independence of all endpoints, i.e. $r = 0$. For D/AP and SiSi, the average of all pairwise absolute correlations was used as described above. The former method still shows similar inflations as presented in Sankoh, Huque (1). Estimating further scaling factors to compensate for the increased FPRs was omitted for SiSi, since the method

**Table S1: False-positive rates of three multiplicity correction approaches.**

| | K \ r | 0.0 | 0.1 | 0.3 | 0.5 | 0.7 | 0.9 |
|---|---|---|---|---|---|---|---|
| Sidak | 2 | 0.0510 | 0.0508 | 0.0468 | 0.0467 | 0.0439 | 0.0407 |
| | 5 | 0.0493 | 0.0472 | 0.0451 | 0.0421 | 0.0357 | 0.0242 |
| | 10 | 0.0492 | 0.0483 | 0.0477 | 0.0392 | 0.0319 | 0.0154 |
| | 100 | 0.0467 | 0.0485 | 0.0433 | 0.0285 | 0.0173 | 0.0051 |
| | 1000 | 0.0491 | 0.0516 | 0.0376 | 0.0228 | 0.0115 | 0.0019 |
| | 10000 | 0.0495 | 0.0458 | 0.0318 | 0.0144 | 0.0047 | 0.0003 |
| D/AP | 2 | 0.0535 | 0.0543 | 0.0565 | 0.0657 | 0.0674 | 0.0703 |
| | 5 | 0.0559 | 0.0570 | 0.0703 | 0.0864 | 0.0933 | 0.0905 |
| | 10 | 0.0581 | 0.0633 | 0.0916 | 0.1056 | 0.1285 | 0.1026 |
| | 100 | 0.0694 | 0.0807 | 0.1351 | 0.1845 | 0.1989 | 0.1582 |
| | 1000 | 0.0842 | 0.1011 | 0.1850 | 0.2604 | 0.2770 | 0.2040 |
| | 10000 | 0.1051 | 0.1291 | 0.2519 | 0.3191 | 0.3363 | 0.2647 |
| SiSi | 2 | 0.0510 | 0.0510 | 0.0480 | 0.0508 | 0.0526 | 0.0576 |
| | 5 | 0.0498 | 0.0479 | 0.0483 | 0.0524 | 0.0540 | 0.0533 |
| | 10 | 0.0495 | 0.0499 | 0.0541 | 0.0517 | 0.0565 | 0.0492 |
| | 100 | 0.0480 | 0.0514 | 0.0531 | 0.0482 | 0.0504 | 0.0451 |
| | 1000 | 0.0503 | 0.0542 | 0.0497 | 0.0469 | 0.0428 | 0.0375 |
| | 10000 | 0.0520 | 0.0508 | 0.0453 | 0.0386 | 0.0330 | 0.0303 |

D/AP was calculated using the average of the absolute correlations between all simultaneously tested endpoints for comparability to the new sine similarity (SiSi) algorithm. Red cells indicate liberal and blue cell conservative settings.

**Table S2: Overview of the psychometric and behavioral data.**

| Score | SCP pre | SCP post | TAU pre | TAU post | p-value |
|---|---|---|---|---|---|
| Emotion Regulation Checklist: emotion regulation subscale | 21.19 ± 3.98 | 23.05 ± 3.23 | 20.45 ± 5.00 | 21.15 ± 3.59 | time: 0.0436 [2] |
| Emotion Regulation Checklist: lability / negativity subscale | 33.71 ± 5.83 | 30.76 ± 5.34 | 32.00 ± 7.06 | 31.65 ± 6.43 | time: 0.0525 [2] ADHD score.: 0.0014 [2] |
| Griffith Empathy Measure: Cognitive empathy subscale | -1.00 ± 9.85 | 5.33 ± 9.80 | 0.60 ± 9.20 | 1.20 ± 8.89 | time: 0.0624 [2] |

Pre- and post-treatment scores are given as mean ± standard deviation. ERC: Emotion Regulation Checklist; ER: emotion regulation subscale; LN: lability/negativity subscale; GEM CE: cognitive empathy subscale of the Griffith Empathy Measure; ADHD: attention-deficit / hyperactivity disorder. [1] two-sample t-test, [2] linear mixed effects model.

stays reasonably close to the nominal significance level (or becomes slightly conservative for a high number of strongly dependent endpoints).

### Supplementary results

P-values of the questionnaire data, RS and brain regulation task results corrected with the Sidak method assuming independence for comparison are provided in Table S2 and Table S3, respectively. A higher conservativity of the Sidak method can be seen, even though the custom algorithm did not yield additional significant findings.

The whole-brain correlation pattern leading to positive relationships between the PerAF clusters and the total displacement measure of movement is shown in the first row of Figure S1. The patterns for ReHo and GFC are provided below for comparison.

**Figure S1: Correlation between resting-state models and SLOMOCO total displacement.**

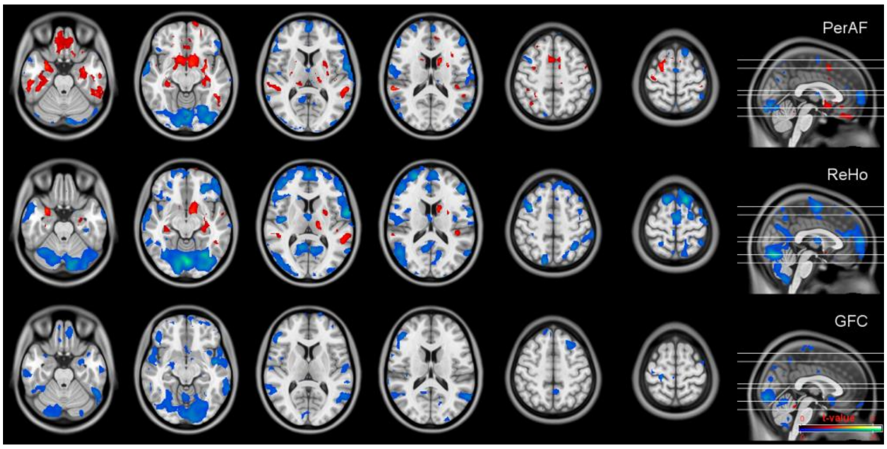


The t-values are thresholded at $p < 0.05$ uncorrected as used in the exploratory parts of the main analyses. PerAF: percent amplitude of fluctuations, ReHo: regional homogeneity, GFC: global functional connectivity.

**Table S3: Results of the whole-brain resting-state analysis and brain regulation task with and without control for movement on group level and Sidak correction.**

| | Model / condition | Contrast | Coordinates x | y | z | Cluster size | p | $p_{corr}$ | Region |
|---|---|---|---|---|---|---|---|---|---|
| Resting-state | PerAF | post > pre | 63 | -34 | -16 | 56 | 0.0481 | 0.1376 | middle / inferior temporal gyrus |
| | PerAF | post > pre | 12 | 29 | 32 | 140 | 0.0003 | 0.0008 | anterior cingulate cortex |
| | PerAF | TAU: post > pre | 60 | -31 | -19 | peak | 0.0433 | 0.1243 | middle / inferior temporal gyrus |
| | PerAF | TAU: post > pre | 57 | -10 | 47 | 96 | 0.0033 | 0.0100 | precentral gyrus |
| | PerAF - total displacement | post > pre | 63 | -34 | -16 | peak | 0.0300 | 0.0872 | middle / inferior temporal gyrus |
| | PerAF - total displacement | post > pre | 63 | -34 | -16 | 56 | 0.0446 | 0.1279 | middle / inferior temporal gyrus |
| | PerAF - total displacement | post > pre | 12 | 29 | 32 | 133 | 0.0003 | 0.0010 | anterior cingulate cortex |
| | PerAF - total displacement | TAU: post > pre | 57 | -10 | 47 | 81 | 0.0073 | 0.0217 | precentral gyrus |
| | PerAF - total displacement | TAU: post > pre | 54 | 23 | 29 | 58 | 0.0385 | 0.1111 | triangular gyrus |
| Brain regulation task | Down | mean > 0 | -18 | -82 | -16 | 543 | 1.8E-9 | 5.5E-9 | primary / secondary visual cortex |
| | Down | mean > 0 | -18 | -82 | -16 | peak | 5.7E-5 | 0.0002 | |
| | Up | mean > 0 | 6 | -79 | -10 | 816 | 2.3E-12 | 6.8E-12 | |
| | Up | mean > 0 | 6 | -79 | -10 | peak | 0.0021 | 0.0064 | |
| | Down | ER > 0 | -39 | -13 | 14 | 141 | 0.0026 | 0.0117 | Rolandic operculum, insula, superior temporal gyrus |
| | Down | ER > 0 | 45 | -4 | -13 | peak | 0.0381 | 0.2950 | insula |
| | Up-Down | LN < 0 | -12 | 35 | 20 | 112 | 0.0066 | 0.0575 | anterior cingulate cortex |
| | Up | ΔGEM cognitive > 0 | 54 | -49 | -22 | 72 | 0.0215 | 0.3263 | inferior temporal gyrus |
| | Up-Down | ΔGEM cognitive > 0 | 51 | -46 | -16 | 157 | 4.2E-4 | 0.0076 | inferior temporal gyrus |

Resting-state: The original results (PerAF) and those additionally corrected for the total displacement (PerAF - total displacement) are provided. Brain regulation task: The mean activation per condition (mean > 0) and the correlation with psychometric score changes (ERC ΔER > 0, ER ΔLN < 0, GEM ΔCE > 0) are shown. All models were corrected for the baseline attention-deficit / hyperactivity disorder (ADHD) score. The coordinates need to be seen as rough orientation only due to a study-specific normalization. The p-values were corrected for the investigated models / contrasts. PerAF: percent amplitude of fluctuation; TAU: treatment as usual; SCP: slow cortical potentials neurofeedback; pre / post: measurement before / after intervention, Up: up-regulation, Down: down-regulation, Up-Down: difference between up- and down-regulation; ERC ΔER: Emotion Regulation Checklist emotion regulation subscale change; ERC ΔLN: Emotion Regulation Checklist lability / negativity subscale change; GEM ΔCE: Griffith Empathy Measure cognitive empathy subscale.
